# Low seropositivity and sub-optimal neutralisation rates in patients fully vaccinated against COVID-19 with B cell malignancies

**DOI:** 10.1101/2021.07.19.21260762

**Authors:** TA Fox, AA Kirkwood, L Enfield, M O’Reilly, S Arulogun, S D’Sa, J O’Nions, J Kavi, E Vitsaras, W Townsend, SO Burns, SH Gohil, K Cwynarski, KJ Thomson, M Noursadeghi, RS Heyderman, T Rampling, KM Ardeshna, LE McCoy, EC Morris

## Abstract

Patients with haematological malignancies are at increased risk of severe disease and death from COVID-19 and are less likely to mount humoral immune responses to COVID-19 vaccination, with the B cell malignancies a particularly high-risk group.

Our COV-VACC study is evaluating the immune response to COVID-19 vaccination in patients with B cell malignancies. Eligible patients were either receiving active treatment or had received treatment within the last 24 months. Patients were vaccinated with either the BNT162b2 (Pfizer-BioNTech) (n=41) or ChAdOx1 nCoV-19 (Oxford-AstraZeneca) (n=14) vaccines. The median age of participants was 60 years (range: 27-82) and 50% were receiving systemic anti-cancer therapy (SACT) at the time of vaccination. This interim analysis from the first 55 participants describes anti-S seropositivity rates, neutralising antibody activity and association with peripheral lymphocyte subsets.

After the first vaccine dose, 36% overall had detectable anti-S antibodies rising to 42% after the second dose. Sera from seropositive patients was assessed for neutralisation activity in vitro. Of the seropositive patients after first dose (n=17), only 41% were able to neutralise SARS-CoV-2 pseudotyped virus with a 50% inhibitory dilution factor (ID50) of >1:50. After two doses (n=21) 57% of the seropositive patients had detectable neutralisation activity (median ID50 of 1:469, range 1:70 – 1:3056). Total blood lymphocyte, CD19, CD4 and CD56 counts were significantly associated with seropositivity. Patients vaccinated more than 6 months after completing therapy were significantly more likely to develop antibodies than those within 6 months of treatment or on active treatment; OR: 5.93 (1.29 – 27.28).

Our data has important implications for patients with B cell malignancies as we demonstrate a disconnect between anti-S seropositivity and virus neutralisation *in vitro* following vaccination against COVID-19.

Urgent consideration should be given to revaccinating patients with B-cell malignancies after completion of anti-cancer treatment as large numbers currently remain at high risk of infection with the increasing transmission of SARS-CoV-2 in many countries.

## Main Text

Patients with haematological malignancies are at increased risk of severe disease and death from COVID-19.^1^ Vaccination is essential to increase population immunity and decrease disease burden. The first COVID-19 vaccines were authorised in the United Kingdom (UK) after phase III trials which showed that both the BNT162b2 (Pfizer-BioNTech) and ChAdOx1 nCoV-19 (Oxford-AstraZeneca) vaccines were highly effective at preventing symptomatic disease and hospitalisation.^2,3^ Real-world vaccine efficacy studies have subsequently shown that both vaccines confer protection against the Delta variant of concern (VOC) (B 1.617.2) which is currently dominant in the UK and elsewhere.^4^

Whilst both vaccines have demonstrated robust immune responses in healthy volunteers, patients with haematological malignancies were excluded from the large immunogenicity and efficacy clinical trials. Emerging data suggests such patients are less likely to mount a humoral immune response to COVID-19 vaccination.^5-7^ Data has shown that patients who have recently received Bruton’s Tyrosine Kinase inhibitors (BTKi) or CD20-directed therapies for B cell malignancies were the least likely to generate SARS-CoV-2S antibodies following vaccination.^8,9^ With the increasing transmission of SARS-CoV-2 in many countries there is an urgent need to understand the risk this poses to vaccinated patients with B cell malignancies.

We report interim results from the first 55 participants recruited to our ongoing COV-VACC study, exploring the immune response to COVID-19 vaccination in patients with B-cell malignancies (South Central Berkshire B Research Ethics Committee and UK Health Research Authority approval IRAS number: 294547). Patients receiving treatment or who had received treatment in the last 24 months for a B cell malignancy and receiving either the BNT162b2 (Pfizer-BioNTech) (n=41) or ChAdOx1 nCoV-19 (Oxford-AstraZeneca) (n=14) vaccines were eligible for recruitment. The median age of participants was 60 years (range: 27-82) and 50% were receiving systemic anti-cancer therapy (SACT) at the time of vaccination. Details of underlying diagnoses and SACT are shown in Figures 1A, B.

**Figure 1.**
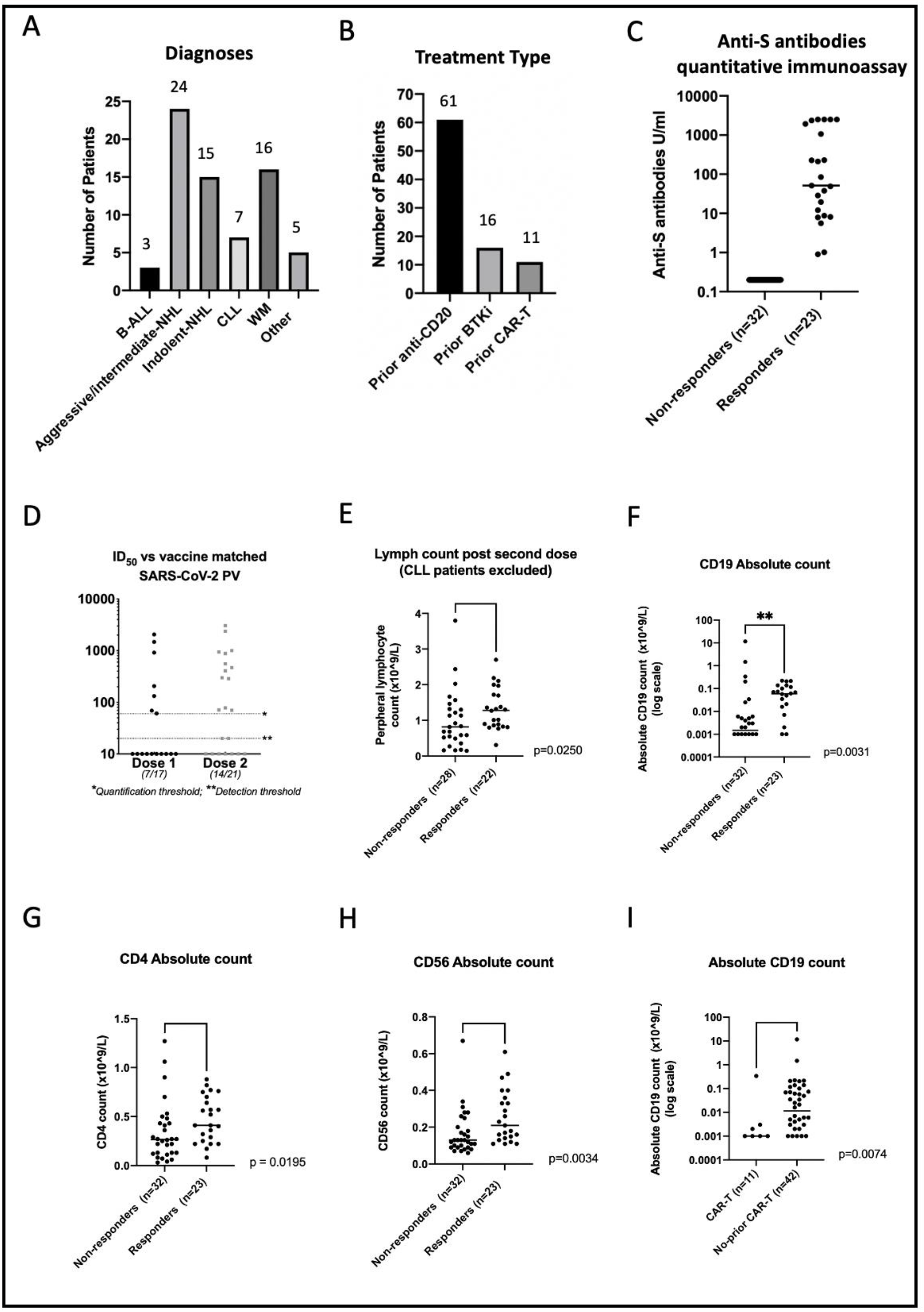
(A) Number of patients by diagnostic group recruited to the study to date (n=70); (B) Number of patients (whole cohort) exposed to common therapeutic modalities; (C) Anti-S antibody levels 1 month post 2^nd^ vaccination quantified by Elecsys Roche Anti-SARS-CoV-2 S assay (Spike); (D) ID50s of serum (from seropositive patients) able to neutralise SARS-CoV-2 pseudotyped virus after first dose (7/17) and second dose (14/21); (E) Peripheral lymphocyte count (excluding CLL patients) in responders (n=22) and non-responders (n=28) post 2^nd^ vaccination (p=0.0250); (F) Peripheral CD19 counts in responders (n=23) and non-responders (n=32) post 2^nd^ vaccination (p=0.031); (G) Peripheral blood CD4 count in responders (n=23) and non-responders (n=32) post 2^nd^ vaccination (p=0.00195); (H) Peripheral blood CD56 count in responders (n=23) and non-responders (n=32) post 2nd vaccination (p=0.0034); (I) Peripheral CD19 count in patients who had received CAR-T therapy (n=11) versus those who had received a different SACT (n=42) (p=0.0074). ‘Responders’ = seropositive with anti-S antibody level >0.4U/ml.

Blood samples were taken prior to vaccination where possible, and 1 month after both first and second vaccine doses. At each time point, a full blood count, and immunoglobulin levels, and enumeration of whole blood lymphocyte subsets (CD3, CD4, CD19, CD56) by flow cytometry (Beckman Coulter (CA, USA) Aquios flow cytometers) were performed. Serum samples were screened for anti-SARS-CoV-2 antibodies using quantitative double-antigen sandwich immunoassays for both the nucleocapsid (N) antigen and the spike (S) protein receptor binding domain (RBD) (both Roche). Samples from participants with detectable anti-S antibodies were then assessed to determine if these antibodies were able to neutralise SARS-CoV-2 infection in vitro using a luciferase encoding lentivirus pseudotyped with the SARS-CoV-2 spike as previously described.^10,11^ Groups were compared using logistic regression, Chi squared/Fisher’s exact tests and Wilcoxon-Mann Whitney tests.

After a single dose of either BNT162b2 (Pfizer-BioNTech) n=41 or ChAdOx1 nCoV-19 (Oxford-AstraZeneca) n=14 vaccine, 36% overall had detectable anti-S antibodies (15/41 Pfizer-BioNTech and 5/14 AZ), and 42% (23/55) after a second dose (Figure 1C). Three participants had serological evidence of previous infection with SARS-CoV-2 (as demonstrated by positive anti-N antibodies).

Where available, sera from seropositive participants after first or second dose were then used to assess neutralisation activity in vitro. Of the seropositive patients after first dose (n=17), just 41% were able to neutralise SARS-CoV-2 pseudotyped virus with a 50% inhibitory dilution factor (ID50) of >1:50. After two doses (n=21) 57% of the seropositive patients had detectable neutralisation activity (median ID50 of 1:469, range 1:70 – 1:3056) (Figure 1D).

Total blood lymphocyte, CD19, CD4, and CD56 counts all showed a significant association with seropositivity (Figures 1E, F, G, H). For a 1 log increase in each lymphocyte subset, the odds of developing antibodies in response to vaccination were 1.32 (95% CI: 1.05 – 1.66, p= 0.013), 2.5 (95% CI: 1.12 – 5.55, p = 0.025) and 4.47 (95% CI: 1.46 – 13.06, p = 0.0008) times higher, respectively for CD19, CD4 and CD56 counts (Table 1). Timing of vaccination in relation to SACT was important (p=0.0126), with participants vaccinated more than 6 months after completing therapy more likely to develop antibodies; OR: 5.33 (1.14 – 24.90). No difference in seropositivity was seen between those vaccinated within 6 months of treatment or on active treatment; OR: 0.68 (0.18 – 2.55). We could not show any impact of therapy type (BTKi or anti-CD20 antibodies), but numbers not treated with CD20 antibodies were very small.

**Table 1.** Logistic regression analysis.

|  | All patients:<br>Anti-S Ab positivity |  |  | Seropositive patients only:<br>neutralising activity |  |  |
| --- | --- | --- | --- | --- | --- | --- |
|  | Seropositive<br>/N | OR (95% CI) | p-value | Neutralising<br>/N | OR<br>(95% CI) | p-value |
| **Lymphocyte subsets** |  |  |  |  |  |  |
| **CD19 (1 log increase)** | 23/54 | 1.32<br>(1.06 – 1.66) | 0.013 | 12/17 | 2.04<br>(0.99 – 4.22) | 0.054 |
| **CD4 (1 log increase)** | 23/53 | 2.50<br>(1.12 – 5.55) | 0.025 | 12/19 | 1.23<br>(0.28 – 5.39) | 0.78 |
| **CD56 (1 log increase)** | 23/53 | 4.47<br>(1.46 – 13.06) | 0.008 | 12/19 | 6.79<br>(0.62 – 73.92) | 0.12 |
| **Treatments** |  |  |  |  |  |  |
| **Rituximab** |  |  |  |  |  |  |
| No | 1/4 | 1.00 | 0.49 | 0/1 | - | 0.37* |
| Yes | 22/51 | 2.28<br>(0.22 – 23.39) |  | 12/18 | - |  |
| **BTKi** |  |  |  |  |  |  |
| No | 17/42 | 1.00 | 0.72 | 11/16 | 1.00 | 0.54 |
| Yes | 6/13 | 1.26<br>(0.36 – 4.41) |  | 2/4 | 0.57<br>(0.05 – 4.67) |  |
| **CAR-T** |  |  |  |  |  |  |
| No | 221/46 | 1.00 | 0.21 | 13/18 | - | 0.37* |
| Yes | 2/9 | 0.34<br>(0.06 – 1.82) |  | 0/1 | - |  |
| **Vaccine time point** |  |  |  |  |  |  |
| On treatment | 9/25 | 1.00 | 0.026 | 3/7 | - | 0.034* |
| Within 6 months of<br>treatment | 5/18 | 0.68<br>(0.18 – 2.55) |  | 2/5 | - |  |
| >6 months from<br>treatment | 9/12 | 5.33<br>(1.14 – 24.90) |  | 7/7 | - |  |

\*OR not estimable, Fisher's exact test used (p = 0.017) for on treatment/within 6 months vs >6 months)

Patients on or within 6 months of treatment had significantly lower CD56 and CD19 counts (p= 0.003 and p= 0.014) and a trend towards lower CD4 (p = 0.11). CAR-T cell therapy recipients had very low rates of seropositivity (2/9, 22.2%) (Table 1). Nine of ten CAR-T therapy patients were within 6 months of infusion.

Seropositive patients could be divided into those whose sera did or did not demonstrate neutralising activity. Neutralising activity was associated with higher median anti-S antibody levels (p=0.0005). Further, both higher CD56 and CD19 counts showed trends towards increased odds of developing neutralising antibodies; OR: 6.79 (0.62 – 73.9), p = 0.12 and 2.04 (0.99 – 4.22), p = 0.054. All seropositive patients (7/7) who were >6 months from treatment had neutralising antibodies compared to 5/12 on or within 6 months of treatment (Fisher’s exact p = 0.017).

This interim analysis adds to a growing body of evidence that immunocompromised patients are less likely to produce robust immune response following COVID-19 vaccination.^5-7,9^ In our cohort 42% had detectable anti-S antibodies following two doses of an approved vaccine compared to 91%-100% in healthy individuals in phase I/II trials.^2,12^ What is striking from the neutralisation data, is that even when seroconversion occurs the protective humoral response may be limited. Just 23% of the cohort (n=56) (57% of seropositive participants) neutralised virus in vitro. Others have shown neutralising antibody levels to be highly predictive of immune protection from symptomatic infection.^13,14^ Our data identifies several factors associated with vaccine response such as peripheral blood lymphocyte, CD19, CD4 and CD56 counts, which if validated in larger cohorts may enable the identification of patients unlikely to respond to vaccination based on their individual immune profile.

This data has important implications for patients with B-cell malignancies and other severely immunocompromised groups, providing further evidence that patients on SACT are less likely to produce antibodies following COVID-19 vaccination.^7^ Firstly, anti-S seropositivity does not necessarily correlate with serum neutralisation and is unlikely predictive of an effective antibody response based on current estimates of correlates of protection.^14^ Urgent validation in larger cohorts is required as many patients with B cell malignancies may remain at high risk of infection, regardless of anti-S antibody status, when community prevalence of COVID-19 is high or increasing. Secondly, all clinically vulnerable patients regardless of vaccination status, should be considered for neutralising monoclonal antibody therapies if they develop COVID-19.^15, 16^

Urgent consideration needs to be given to provision of booster doses or full revaccination to this group of patients, particularly if they have been vaccinated within 6 months of active therapy. A third vaccination dose has been shown to be beneficial in solid organ transplant recipients ^17^, although low CD19 and CD4 counts were also associated with poor responses. The correlation between peripheral blood lymphocyte, CD19, CD4 and CD56 counts suggest that booster doses or vaccination may be most effective if given when an individual has recovered lymphocytes and are at least 6 months following SACT.

This interim analysis is limited by cohort size and heterogeneity. However, we demonstrate a disconnect between seropositivity and virus neutralisation *in vitro*, following vaccination against COVID-19.

## Data Availability

No supplementary data submitted

## Acknowledgements

This study is supported by the NIHR UCLH Biomedical Research Centre and Blood Cancer UK. RSH is a NIHR Senior Investigator. The views expressed in this publication are those of the author(s) and not necessarily those of the NIHR or the Department of Health and Social Care.

